# Reward Dysfunction in General and Specific Psychopathology in Children and Adults

**DOI:** 10.1101/2021.10.13.21264963

**Authors:** Ankita Saxena, Catharina A. Hartman, Steven D. Blatt, Wanda P. Fremont, Stephen J. Glatt, Stephen V. Faraone, Yanli Zhang-James

**Affiliations:** Departments of Psychiatry and Behavioral Sciences, State University of New York – Upstate Medical University, Syracuse, New York, USA; Department of Psychiatry, University of Groningen, University Medical Center Groningen, Groningen, The Netherlands; Department of Pediatrics, SUNY Upstate Medical University, Syracuse, New York, USA

**Author notes:** **Contact Information for Corresponding Author:** Yanli Zhang-James, PhD.

**Keywords:** Psychopathology, P-factor, Neuropsychiatric disorders, Disorder comorbidity, Reward Motivation, Reward Valuation, Reward Responsiveness

## Abstract

**Background:** Reward dysfunction has been implicated in many psychiatric disorders such as attention-deficit/hyperactivity disorders, depression, and substance use disorders. However, psychiatric comorbidities are common, and the specificity of reward dysfunction to individual psychopathologies is unknown. The objective of this study was to evaluate the association between reward functioning and specific or general psychopathologies.

**Methods:** 1044 adults and their 1215 children (ages 6-12) completed various measures of the Positive Valence System domain from the Research Domain Criteria (RDoC), which included the Delayed and Probability Discounting Tasks, Energy Expenditure for Reward Task, and Iowa Gambling Task. Children also completed the Experienced Pleasure Scale for children, while adults completed the Temporal Experience of Pleasure Scale and the Behavioral Activation System tasks. Psychopathology was assessed using the Child Behavior Checklist (CBCL) for children and the Adult Self Report (ASR) for parents.

**Results:** One general factor identified *via* principal factors factor analysis explained the majority of variance in psychopathology in both groups. Reward measures in both adults and children were significantly associated with general psychopathology as well as most specific psychopathologies. Some associations between reward and psychopathology did not hold following removal of general psychopathology; nonetheless, certain reward constructs were uniquely associated with specific disorder problems but not general psychopathology.

**Conclusion:** Disorder problem specific associations with reward functioning can be identified after removal of comorbidity. General propensity toward psychopathology is significantly but not uniquely correlated with reward dysfunction. Altogether, this may have broader implications for future study of the role of reward in disease pathogenesis.

## INTRODUCTION

Although psychiatric disorders are delineated as categories in the Diagnostic and Statistical Manual of Mental Disorders (DSM), comorbidity among disorders is common (Caron & Rutter, 1991). Consequently, these disorders have often been grouped into higher-order factors such as “internalizing” or “externalizing” disorders. However, despite this classification, significant covariation continues to exist between these dimensions, with both clinical and genetic studies implicating a single factor, the *P-factor*, that reflects an individual’s propensity toward developing any psychopathology (Caspi et al., 2014; Lahey, Zald, Hakes, Krueger, & Rathouz, 2014; Selzam, Coleman, Caspi, Moffitt, & Plomin, 2018; Smoller et al., 2019; Waldman, Poore, van Hulle, Rathouz, & Lahey, 2016). Regarded as analogous to the *g* of general intelligence, the presence of the *P-factor* has motivated studies of nonspecific and specific risk factors of psychopathology (Jensen, 1993; Spearman, 1904).

Altered reward functioning has been previously linked with different individual psychiatric disorders. Abnormal delayed discounting has been identified in patients with attention-deficit/hyperactivity disorder (ADHD), addictive disorders, major depressive disorder (MDD), and other conditions (Amlung et al., 2019; Amlung, Vedelago, Acker, Balodis, & MacKillop, 2017; de Castro Paiva, de Souza Costa, Malloy-Diniz, Marques de Miranda, & Jardim de Paula, 2019). Substance abuse disorders (SUDs) and MDD have been associated with anhedonia and increased risky decision-making, as measured by the Iowa Gambling Task (IGT) (de Siqueira et al., 2018; Kovács, Richman, Janka, Maraz, & Andó, 2017; Rizvi, Lambert, & Kennedy, 2018). Additionally, increased impulsive decision-making is found in children with ADHD, though results have been mixed for adults with ADHD (Groen, Gaastra, Lewis-Evans, & Tucha, 2013). In addition to a heightened incidence of anhedonia, risky behavior, and reduced effort-based decision making, fMRI studies have shown aberrant reward processing in MDD patients (Murray, Waller, & Hyde, 2018). Finally, altered neural reactivity to reward has been observed in individuals exhibiting antisocial behavior (Plichta & Scheres, 2014).

The relationship between reward and general psychopathology is not well studied. Understanding the degree to which abnormal reward valuation or responsiveness are independently associated with *P-factor* and specific psychopathologies will enhance our understanding of the link between reward dysfunction and psychiatric illness. We hypothesized that both the *P-factor* and specific disorder problem severity would be independently associated with reward functioning but did not have a directional hypothesis about their relative contributions to this association. In our analysis, we extracted *P* via factor analysis and derived psychopathology specific scores; we then tested the association between reward measures and these measures of general and specific psychopathology.

## METHODS

### Study Cohort

Child probands in the age range of 6-12 years were recruited such that approximately half would have high and half low psychopathology scores. For this, participants were recruited from specific settings. To capture participants with higher psychopathology scores, children receiving clinical services *via* the Child and Adolescent Psychiatry Clinic at SUNY Upstate Medical University, or through community private practice clinicians, were invited to participate. Children with lower psychopathology scores were recruited from community events throughout the greater Syracuse area. After rapport was developed with the proband and their family, they were presented with the opportunity to participate in the study. Adults and children who had sensorimotor disabilities, diagnosed neurological disease, a history of head trauma with a documented loss of consciousness exceeding 10 minutes, an uncontrolled medical condition, a history of psychotropic medication utilization, or lack of comprehension of the English language, were excluded from the study. In addition, adopted children as well as adults who could not independently complete the tasks of the study, and pregnant women or those who gave birth up to 6 months prior to the study visit, were not retained as participants. Potential participants who had an estimated intelligence quotient (IQ) below 80, as computed *via* the vocabulary and abstraction subtests of the Shipley-2, were excluded from the study (Shipley, 1940). Parents older than 59 were excluded to minimize effects of possible cognitive decline. Parents provided informed consent and children assented to the study.

#### Measures

The study visit lasted for approximately three hours, with participants completing several tasks and behavioral assessments.

##### Adult Self Report (ASR)

Psychopathology in adult participants was assessed *via* the ASR, a self-reported, 126-item questionnaire, employed for ages 18-59 (Achenbach & Rescorla, 2003). It is a broadly utilized instrument that evaluates psychopathology, substance use, and adaptive functioning (Achenbach & Rescorla, 2003). T-scores for symptoms of six DSM disorders (depressive disorders, anxiety disorders, somatic problems, ADHD, avoidant personality, and antisocial personality) were computed *via* the ASR (Achenbach, Bernstein, & Dumenci, 2005). It also provides subscales for symptoms of substance abuse (tobacco, alcohol, recreational drugs), as well as a composite total for substance abuse (Achenbach & Rescorla, 2003). In addition, T-score scales are available to describe symptoms of obsessive-compulsive problems (OCP), sluggish cognitive tempo (SCT), as well as stress problems and total problems and measures of internalizing and externalizing behavior (Achenbach & Rescorla, 2003). In this study, T-scores from individual substance abuse subscales, all DSM scales, and OCP and SCT were approximated as measures of problems linked to specific psychopathologies (Achenbach & Rescorla, 2003).

##### Child Behavior Checklist (CBCL)

Psychopathology in children participating in the study was assessed *via* the CBCL, a 113-item, parent-reported questionnaire (Achenbach & Edelbrock, 1991; Ivanova et al., 2007) It assesses behavioral and emotional problems in children from ages 6 to 18 (Ivanova et al., 2007). Six DSM scales are also available for the CBCL (affective problems, anxiety problems, somatic problems, ADHD, oppositional defiant problems and conduct problems), as are scales for SCT and obsessive compulsive problems (OCP) (Nakamura, Ebesutani, Bernstein, & Chorpita, 2009). T-scores from these scales were employed in the study.

##### Experienced Pleasure Scale for Children (EPSC)

was utilized to measure initial responsiveness to reward in children. It was derived from Kazdin’s original 39-item, self-report inventory, the Pleasure Scale for Children (PSC), that evaluates anhedonia in children of school age (Kazdin, 1989). Items are reflective of three categories of anhedonia: physical anhedonia or physical pleasures, social anhedonia or pleasures associated with interaction with others, and other reward related activities such as interests and achievements. For each item, children rank their enjoyment of an activity as either making them, “very happy” (3), “happy,” (2), or “wouldn’t matter,” (1). Children who score highly on anhedonia *via* the PSC show reduced reward seeking activity and reduced affect in response to rewarding events (Kazdin, 1989). For the EPSC, participants were first asked to answer either Yes/No as to whether they had personally experienced each scenario, and then directed to rank their enjoyment of scenarios that they had undergone. A mean score was computed for each participant based on their ratings for the items that they endorsed experiencing.

##### Temporal Experience of Pleasure (TEPS)

is an 18-item self-report measure used in adults to assess anticipatory and consummatory pleasure (Gard, Gard, Kring, & John, 2006). 10 items reflect anticipatory pleasure, e.g. pleasure felt in anticipation of a positive stimulus, and eight items consummatory pleasure, which is defined as pleasure felt, “in-the-moment,” in response to a stimulus (Gard et al., 2006). Participants responded to each item on a six-point Likert scale, where 1= very false for me and 6 signified very true for me; item scores were then summed and utilized to generate a composite score. Only the consummatory subscale was obtained in this study as a measure of initial responsiveness to reward attainment. The subscale was sum-scored with scores ranging from 8 to 48; higher scores indicated greater consummatory pleasure and initial responsiveness to reward attainment.

##### Iowa Gambling Task (IGT)

is a measure of risky decision-making (Cauffman et al., 2010; Garon, Moore, & Waschbusch, 2006). Participants are initially given $2000 in play money and are directed to maximize profit through the course of 100 trials by selecting cards from decks A, B, C, or D. Individual cards can yield either a gain or a loss, but repeated selection from decks results in differential outcomes. Although cards from “disadvantageous” decks A and B can yield greater immediate returns, long-term selection results in greater losses, while cards from “advantageous” decks C and D offer smaller immediate gains but better long-term outcomes. At the same time, decks A and C offer frequent losses, whereas decks B and D offer infrequent ones. Consequently, the IGT evaluates the participant’s ability to learn *via* trial and error to select advantageous decks *vs*. disadvantageous ones for a reward. Two summary measures from the task were included in our analysis as measures of reward responsiveness: individual IGT net earnings (IGT-NE) and total latency, evaluated by the amount of time it took to identify advantageous decks (IGTL) (Bull, Tippett, & Addis, 2015).

##### *Effort Expenditure for Rewards Task* (EEfRT)

is a measure of effort-based decision making (Treadway, Buckholtz, Schwartzman, Lambert, & Zald, 2009). In the task, participants complete multiple trials where they choose between the options of completing a “hard task” or an “easy task” to receive a monetary reward. In the easy-task, participants must press a keyboard button with the index finger of their dominant hand 30 times within 7 seconds for an assigned reward value of $1.00. For the hard-task, individuals must complete 100 button presses with the little finger of their non-dominant hand within 21 seconds; the possible reward varied between $1.24 and $4.30. Rewards were not guaranteed upon trial completion; some trials were ‘no-wins’, while others were ‘wins’ that resulted in earnings. Exclusively in the adult version of the task subjects were given the probability of receiving a reward upon trial completion; probabilities were ‘high’ (88%), ‘medium’ (50%), or ‘low’ (12%). Probability levels were uniform for the trial, regardless of task difficulty, and equal proportions of each probability level were utilized through the experiment. Trials were presented in a randomized order to all subjects. Two analysis variables were derived from this measure: individual effort expenditure defined as the percentage of trials for which the participant selected the hard-task (Effort Expenditure for Rewards Choice total, EERCT), and the Effort Expenditure for Rewards Beta coefficient (EERCB), which indexes reward sensitivity (Nguyen et al., 2019). EERCB reflects the standardized beta-weight values from a logistic regression of reward value (independent variable) and hard task choice (dependent variable) (Nguyen et al., 2019).

##### Delayed Discounting Task (DDT) and Probability Discounting Task (PDT)

are measures to assess reward valuation(Richards, Zhang, Mitchell, & de Wit, 1999; Scheres, Tontsch, Thoeny, & Kaczkurkin, 2010). For the DDT, in a series of trials, participants choose between receiving a large reward ($10 for adults, $1 for children) after delays of varying lengths (1 to 365 days for adults, 5-60 seconds for children), or receiving a smaller reward immediately ($2 for adults, $0.20 for children). For each delay, the immediate reward offered was modulated based on previous responses until the subject elected to receive the reward immediately rather than wait to achieve the maximum reward. The minimum reward amount for which subjects did not show a preference between immediate receipt as opposed to waiting for the specified delay for the maximum reward was deemed the indifference value. PDT trials were carried out in between DDT trials for nine-year old children and adults. For these, participants were given a choice between receipt of a large reward at varying levels of probability (25, 50, 75, and 90%) or a smaller, guaranteed amount (100% probability). As with the DDT, the guaranteed amount was increased or decreased in value based on previous responses until an indifference value was obtained.

##### Behavioral Activation System (BAS)

is a 17-item self-inventory that assesses an adult’s sensitivity to approach motivation (Carver & White, 1994). Approach motivation is assessed in three areas, with four items evaluating drive, four, fun-seeking, and five, reward-responsiveness. Each item is scored on a 4-point Likert scale, with responses including: (1) very true for me; (2) somewhat true for me; (3) somewhat false for me; and (4) very false for me. For this study, the analytic variables that were obtained included the individual sum-scores for the drive (BASD) and reward responsiveness (BASR) scales.

#### Statistical Analyses

Statistical analyses were conducted with Stata 15.1. We adjusted for age, sex, race, and ethnicity (*via* dummy variable coding) through ordinary least squares regression. To derive the *P-factor*, a bifactor confirmatory factor analysis, as used in previous studies, was attempted (Caspi et al., 2014). In addition, principal factors factor analysis (PFFA) was conducted using T-scores for the psychopathologies selected for each age group. To evaluate the role of item overlap between the DSM scales and SCT and OCP (4 items per age group), PFFA to identify the *P-factor* was also done exclusively with DSM scales. After the loading matrix was computed, factors with eigenvalues greater than 1 were identified, and then the matrix underwent an orthogonal varimax rotation. The factor with the highest eigenvalue, i.e., Factor 1, was designated the *P-factor*.

Based on this, scores for Factor 1 were assigned to each individual as their *P-factor* values. In addition, the loadings for each individual disorder on Factor 1 were obtained; these values were considered representative of how much variance within each disorder could be explained by the general factor of psychopathology.

To obtain T-score values representative of disorder-specific pathology, T-scores for individual disorders were linearly regressed against *P-factor* scores. The residuals computed were regarded as disorder problem specific T-scores. To identify which subset of reward variables were best suited for predicting the *P-factor* and disorder problem specific T-scores, the Furnival-Wilson leaps-and-bounds algorithm using logarithmic likelihoods was applied through the Stata function ‘GVSelect.’ The variables in the models that yielded low Bayesian and Akaike information criterions were used in linear regression.

The relationships between the reward measures, the *P-factor*, and disorder-specific T-scores were evaluated by linear regression analysis. The *F*-test of the overall significance was used to assess whether the null hypothesis could be rejected and that the model with the proposed variables provided superior fit to a constant-only model. In order to account for the increased risk of type-I error due to multiple tests, the Benjamini-Hochberg method was used to adjust calculated *p*-values (Chen, Feng, & Yi, 2017). Nine tests were conducted for the child data and 12 for the adult data. To determine statistical significance, we used a 5% False Discovery Rate. Two-tailed test *p*-values were computed for each regression coefficient and similarly compared to the alpha.

To evaluate the univariate relationship between reward measures and disorders, Pearson’s correlation coefficients were computed between reward measures and disorder problem severity. Correction for multiple testing was addressed as described for the regression analysis. 17 and 24 tests were conducted for child and adult data, respectively.

## RESULTS

### Population

1215 children, with an average age of 9 years (S.D. = 2.2) and 1044 parents between the ages of 23 and 59 years (mean = 37 years, S.D. = 7.2) made up the study population. In comparison to the general population, our study sample was enriched for psychopathology by design, with 45% of parents and 41% of children reporting a psychiatric history, as defined by having experiences of previously seeking mental healthcare for emotional or behavioral issues.

The sample had nearly equal numbers of male and female juvenile participants (51% to 49%, respectively), albeit females were overrepresented in the parental population, (69% female, 31% male). 55% of children and 65% of parents identified as White, 25% of children and 24% of parents as Black, and 20% of children and 11% of adults as ‘other’ or multiple races. In addition, approximately 11% of children and 7% of parents identified as Hispanic. Altogether, the dataset consisted of 770 different families with average size of 2.93 individuals.

### Factor Analyses of Psychopathology

The Bifactor Confirmatory Factor Analysis failed to converge; thus, PFFA was alternatively used. PFFA for both child and adult disorders yielded only one factor, i.e., the *P-factor*, with an eigenvalue greater than 1. In children, *P* explained 54% of variance, and in adults, over 95%. Overall, anxiety problems loaded the highest on the *P-factor* in children, with externalizing disorders such as oppositional defiant disorder and conduct disorder loading the lowest (Figure 1). In adults, all non-substance use-related psychopathologies loaded substantially on *P*, with substance pathologies showing very low loadings (Figure 2).

**Figure 1:**
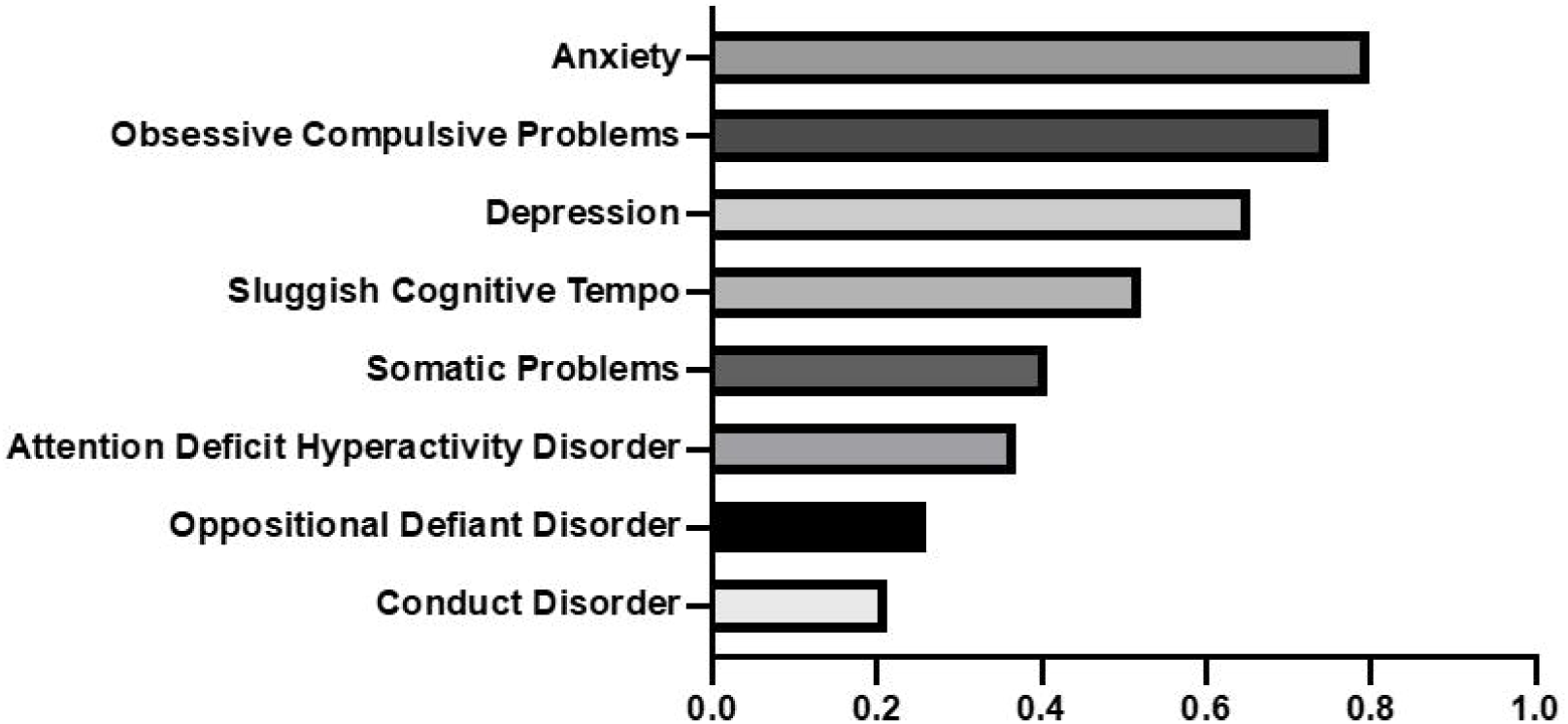
Loadings for specific disorders on the common factor, ‘P’, in children.

**Figure 2:**
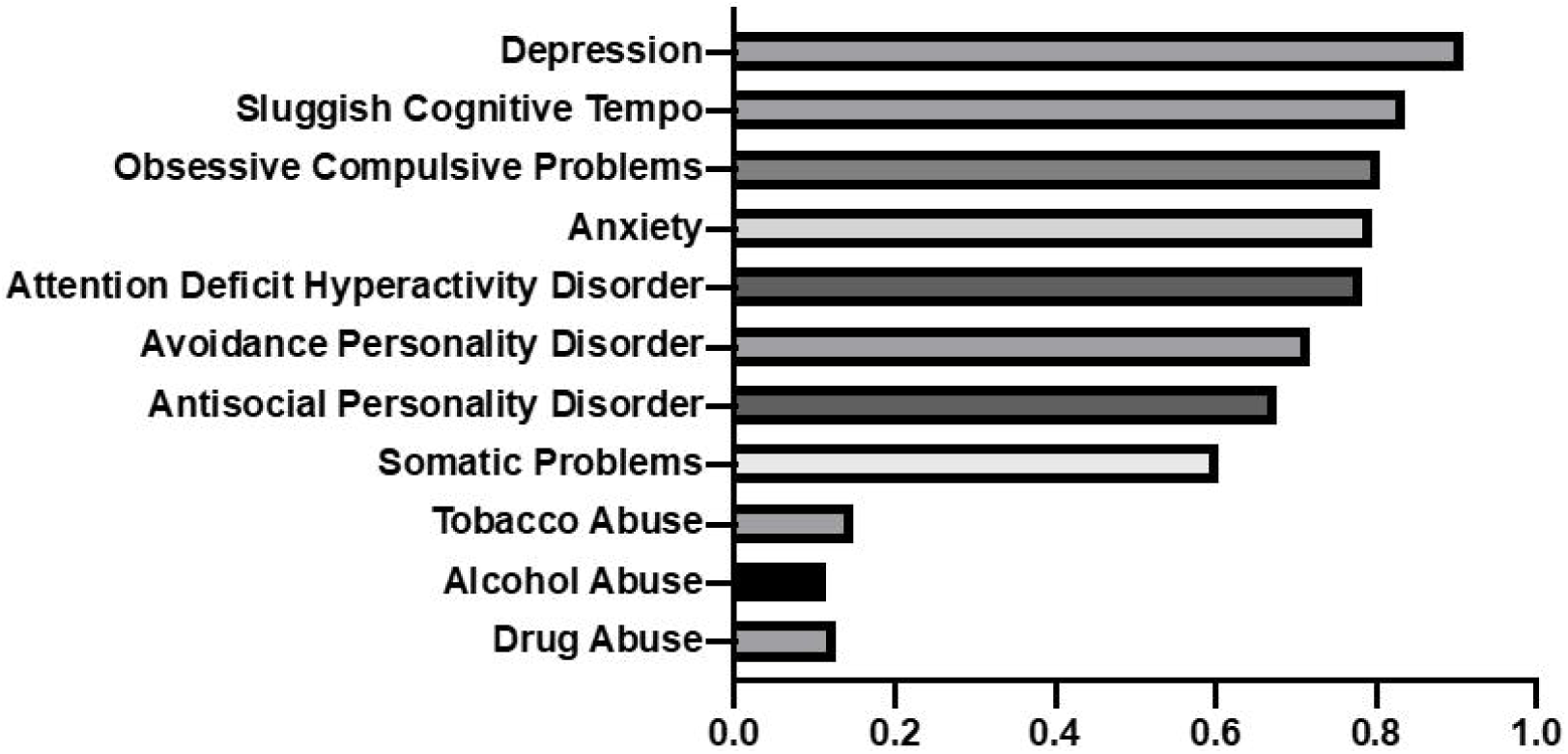
Loadings for specific disorders on the common factor, ‘P’, in Adults.

PFFA on datasets with OCP and SCT removed similarly yielded 1 general factor, with it accounting for ∼67% of variance in children and ∼99% in adults. In both age groups, loadings on Factor 1 were increased; however, the overall patterns were the same.

### Reward Measures and Psychopathology in Children

Five of the seven measures of reward significantly correlated with the *P-factor* and disorder-specific psychopathology scores: EERCT, PDT, DDT, IGT-NE and EERCB (see Table 1). Table 1 shows that nearly all psychopathologies, except for OCP and Somatic problems, were significantly associated with reward measures; however, most disorder problems were linked to only a few reward measures. Table 1 shows the correlations between reward measures and psychopathology; empty table entries indicate that the correlation was not significant. One reward scale (DDT) was associated with both general psychopathology and specific psychopathologies, while the rest were uniquely associated with disorder problems (see Table 1).

**Table 1:** Correlation of Reward Functioning with General and Specific Psychopathologies in Children

| Dependent Variable | Independent Variables |  |  |  |  |  |  |  |  |
| --- | --- | --- | --- | --- | --- | --- | --- | --- | --- |
|  | Pleasure Scale | Energy Expenditure for Reward Choice total | Iowa Gambling Total Net Earnings | Probability Discounting Task | Delayed Discounting Task | Energy Expenditure for Reward Choice Beta coefficient | Iowa Gambling Total latency | Model F-statistic | Model P-value |
| P-factor |  |  |  |  | 0.061 (0.028) |  |  | 5.62 | 0.023 |
| ADHD |  | -0.074 (0.0097) |  |  |  |  |  | 8.33 | 0.0060 |
| Depression |  | -0.068 (0.015) | -0.057 (0.034) | -0.068 (0.015) |  |  |  | 6.28 | 0.00090 |
| Anxiety |  |  |  |  | -0.10 (<0.001) | -0.086 (0.0031) |  | 13.2 | <0.0001 |
| ODD |  |  | -0.052 (0.050) |  | -0.074 (0.0097) |  |  | 6.39 | 0.0031 |
| OCS |  |  |  | -0.047 (0.073) |  |  |  | 3.30 | 0.078 |
| Somatic Problems |  | -0.031 (0.23) |  |  |  |  |  | 1.45 | 0.23 |
| CD |  | -0.065 (0.020) | -0.052 (0.050) | -0.13 (<0.001) | -0.097 (0.0011) |  |  | 10.2 | <0.0001 |
| Sluggish Cog. Tempo |  | -0.086 (0.0031) |  | -0.057 (0.034) |  |  |  | 7.80 | 0.00090 |

Note: ODD = Oppositional Defiant Disorder, OCP = Obsessive Compulsive Symptoms, CD = Conduct Disorder, SCT = Sluggish Cognitive Tempo. In all columns absent the last two, the first number listed is the Pearson's correlation coefficient, while the value in the parentheses is the coefficient P-value. FDR correction was applied to values in the Model P-value column, and to coefficients.

### Reward Measures and Psychopathology in Adults

In adults, scores from nine measures (TEPS, BASR, BASD, EERCT, EERCB, IGT-NE, IGTL, PDT and DDT) correlated significantly with general and specific psychopathologies; of the nine, one, EERCT, was significantly associated with the *P-factor* and disorder, while the remainder were unique to disorders (see Table 2). Apart from ADHD, all specific psychopathologies were significantly associated with one or more measure of reward. As with Table 1, table entries for reward measures correlating to specific psychopathologies show their Pearson’s correlation coefficient values and *p*-values; empty entries mean no significant relationship was found. For disorders where no relationship was found, the best performing measure was included. These findings in adults differ from the pattern seen in children, where ADHD problem severity was reward-associated but SCT, OCP, and somatic problems were not.

**Table 2:**
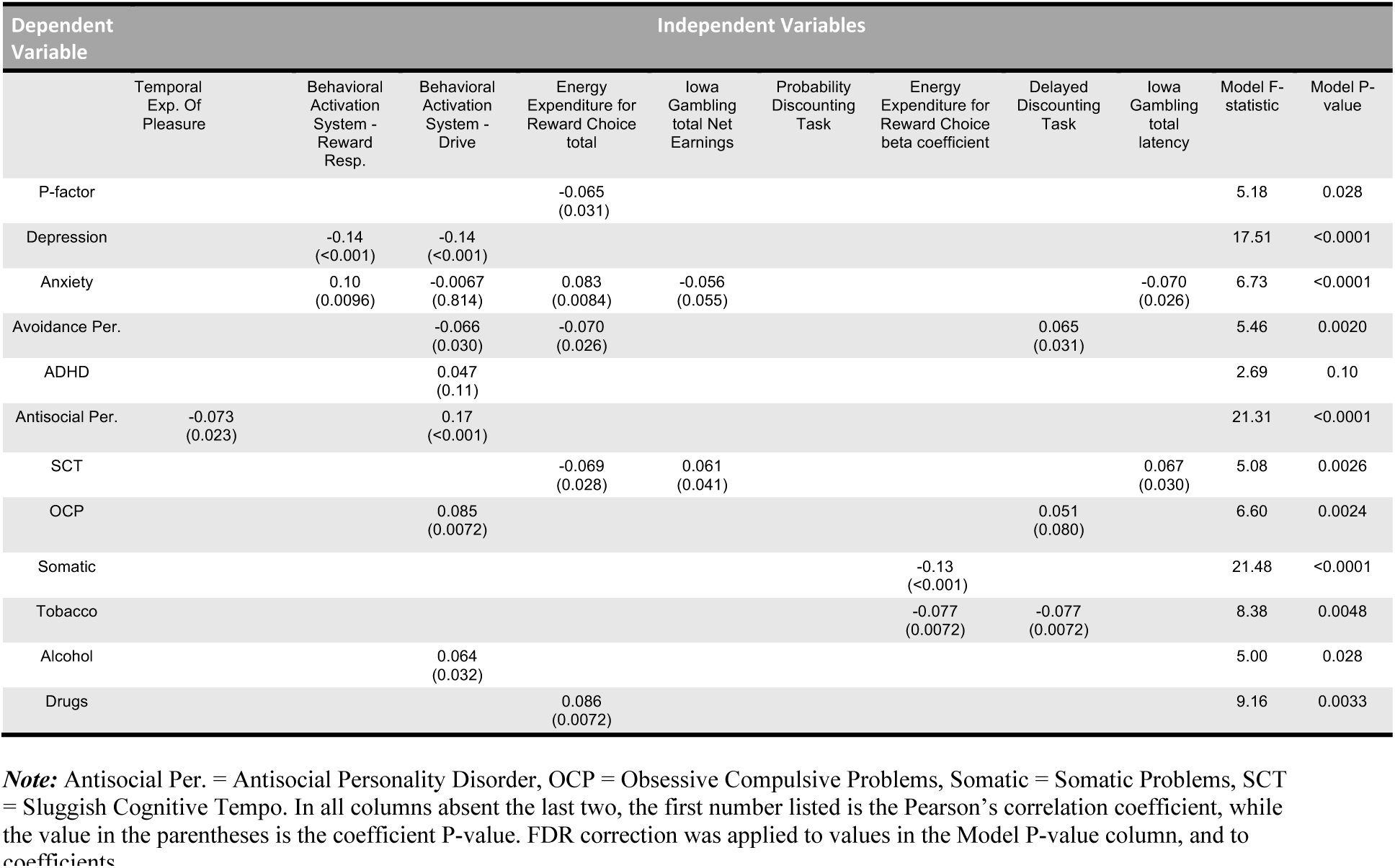
Correlation of Reward Functioning with General and Specific Psychopathologies in Adults:

#### Univariate Correlation Analysis

In both age groups, most reward measures found to be significantly associated with general and specific psychopathologies via regression also yielded statistically significant Pearson’s correlation coefficients. In children, the exceptions were: IGT-NE (ODD, CD), while in adults they were IGT-NE(Anxiety), and DDT(SCT). Overall, correlation coefficients were low, with none exceeding 0.2.

## DISCUSSION

We identified a general factor of psychopathology, *P*, that describes an individual’s overall propensity for psychopathology, by extracting the common factor between multiple disorders through PFFA. This enabled us to identify associations between measures of reward and general psychopathology, as well as specific psychopathologies.

For both children and adults, most variance in the psychopathologies assessed was explained by the *P-factor*, although the amount of variance explained by the *P-factor* in both groups differed greatly. The ∼54% seen in children aligns with earlier findings but the 95% variance explained by the adult *P-factor* is substantially more than in previous reports (Allegrini et al., 2020; Caspi et al., 2014; Selzam et al., 2018). The large percentage of variance accounted for by the *P-factor* suggests that much of a presumed specific psychopathology score can be attributed to general psychopathology, rather than being regarded as reflective of disorder-specific problem severity. Simultaneously, this may have occurred because the CBCL and ASR were not designed with the intent to maximally differentiate among specific domains of psychopathology, allowing for the dominance of a general factor (Achenbach et al., 2005; Hartman, 2021; Hartman et al., 1999; Moore et al., 2020). Importantly, removal of the SCT and OCP resulted in the *P-factor* explaining more variance than before in both age groups and increased loadings for all disorders. This suggests that item overlap did not falsely bolster *P*’s encapsulation of variance, and that scale inclusion slightly improved analysis dimensionality.

Previous work has also reported the existence of lower-order orthogonal factors, such as “internalizing” and “externalizing” disorder groupings; however, none could be identified here (Caspi et al., 2014; Lahey et al., 2014; Martel et al., 2017; Selzam et al., 2018; Waldman et al., 2016). These results may be due to the fact that previous studies incorporated different disorders from different instruments in their analyses. Additionally, earlier studies used multiple time points to derive *P*; repeated measurement of disorder-specific problem severity allows for better differentiation of specific factors due to reduction of state influences and noise. Also, study populations differed – it is conceivable that our sampling of children who were healthy and who had increased problems led to the strong dominance of a single factor, although the fact that the single factor was more dominant in the parent data speaks against this. Factor structure results may thus differ because of the aforementioned characteristics as well as the fact that bifactor CFAs inherently assume an additional structure with specific factors, while PFFA seeks to maximize explanation of variance. Nonetheless, the low dimensionality in our data may very well be the reason why the bifactor CFA did not converge in the first place. Importantly, and in line with all of this, individual disorder loadings on our *P-factor* did not diverge substantially from those found in single factor CFA models (Caspi et al., 2014). Interestingly, in children, “internalizing” problem severity, e.g. Depression, Anxiety, or OCP, had higher loadings on Factor 1 than externalizing disorder problem severity (ADHD, CD, or ODD). Thus, the PFFA derived factor, or *P-factor*, substantially captured aspects of internalizing problems compared with externalizing; this would affect the structure of any subsequent factors extracted. Furthermore, the substantial accounting of variance in both age groups by the *P-factor* inherently limits the explainability of subsequent factors.

The *P-factor* has been previously associated with various measures, including IQ, executive function, and memory (Caspi et al., 2014). In tandem, substantial work has been done on identifying linkages between different measures evaluating reward valuation (BAS, DDT, PDT), reward prediction error, responsiveness to reward attainment (TEPS, PSC, IGT), and reward motivation (EEfRT)) and various psychopathologies. This study extends existing literature in three ways: first, it identifies aberrant reward processing associated with general psychopathology in two age groups; second, it shows that some previously identified disorder-reward dysfunction correlations can be explained by the *P-factor*; third, it identifies areas of reward processing that appear to be disorder-specific. In children, *P* was significantly associated with reduced reward valuation, and with low reward motivation in adults. Both aspects of reward have been previously associated with aberrant psychiatric functioning; steep delayed discounting, identified in children, has been associated with several disorders, including ADHD, while attenuated motivation has been linked to psychiatric symptoms such as lethargy and anergy (Amlung et al., 2019; de Castro Paiva et al., 2019; Rizvi et al., 2018). In turn, these symptoms have been observed in various psychopathologies such as depression or schizophrenia (Rizvi et al., 2018; Trøstheim et al., 2020).

We found that some prior reports of disorder-specific associations could not be replicated after removing their common variance with the *P-factor*. This suggests that aspects of dysfunctional reward functioning previously associated with disorders were likely not specific for the disorder, but rather a consequence of comorbidity. For example, previously, steep delayed discounting was found to be linked with ADHD in children but failed to correlate with ADHD problem severity following removal of the general factor (de Castro Paiva et al., 2019). Most reward measures that had previously been associated with individual disorders were neither linked to the *P-factor*, nor to specific psychopathology T-scores. However, certain psychopathologies maintained previous associations following removal of general psychopathology. For example, antisocial behavior has been strongly linked with increased BAS activity and the same finding was observed following *P* extraction (Hoppenbrouwers, Neumann, Lewis, & Johansson, 2015; Murray et al., 2018). Simultaneously, SUDs have been associated with delayed discounting in the literature, but after *P-factor* removal, only Tobacco UD remained associated (Amlung et al., 2017).

However, even following removal of general psychopathology, most specific psychopathologies in both children and adults were associated here with unique reward measures that were not shared with the *P-factor*. This indicates specific areas of reward dysfunction particularly relevant to individual diseases. For instance, while ODD and *P* are both linked with increased delayed discounting, a measure of altered reward valuation, ODD is also negatively correlated with IGT-NE, a measure reflective of reduced reward responsiveness. Apart from a few disorders that did not correlate with any measures, this allows us to differentiate between mechanisms pertinent to a certain psychopathology versus a broader propensity toward general disease development. Altogether, this finding is relevant because in further studies of reward functioning and disease, it may be of interest to prioritize specific aspects of reward function relevant to unique disorders. Furthermore, understanding the interplay between general psychopathology and these disorder-specific aspects in the context of reward dysfunction may provide mechanistic insight.

Importantly, although many significant associations were found, univariate correlations between reward measures and psychopathologies (see Table 1, Table 2) were low. This suggests that while altered reward functioning is significantly linked with disorders, the magnitude of association is small. This finding is consistent with the understanding that most psychopathologies have multifactorial etiologies; other aspects such as social and cognitive functioning may account for the unobserved associations (Hess, Radonjić, Patak, Glatt, & Faraone, 2020; Radonjić et al., 2021; Zaso, Maisto, Glatt, Hess, & Park, 2020).

Our work must be interpreted in the context of several limitations. First, sampling for our study was enriched for psychopathology, as opposed to a population sample, and the measurement instruments used only evaluated a select group of disorders. Moreover, our population was not representative of severe psychopathology, as individuals were community-dwelling. Further study with inclusion of a greater variety of disorders and conditions that are not assessed via the ASR or CBCL, such as schizophrenia, gambling disorder, or autism spectrum disorder, as well as application of this analysis to different populations would enhance characterization of *P* and associated factors. Adding more disorders and more differentiated measures, possibly ones less explained by general psychopathology, would introduce greater population variance and affect computation of the *P-factor* and downstream calculations. Our failure to replicate previous work may also be due to our study using indirect, continuous measures of psychopathology *via* ASR and CBCL T-scores rather than direct clinical diagnoses. Also, parents were the sole raters for themselves and their children; this dependency in the data opens the possibility for informant biases. Finally, the population demographics of participants also limit the generalizability of this study. Women are over-represented in this sample, and non-Black/African-American minorities are under-represented – additional study with a more balanced population could lead to different findings.

In summary, our work shows relationships between constructs of reward and general psychopathology in both children and adults, implicating different constructs across the two age groups. It also shows that some reward constructs differ in the degree to which they are associated with the *P-factor* and with individual psychopathologies. By identifying general and disorder-specific associations with reward functions, we may be able to separate out reward functions that play a role in the overall predisposition to psychopathology and those that steer adults or children toward a specific trajectory. Altogether, this study advances knowledge about the nature of *P* and suggests that reward mechanisms may explain the emergence of some psychopathologies and their comorbidities.

## Data Availability

All data produced in the present study are available upon reasonable request to the authors and institutional approval

## Acknowledgements

This project has received funding from the European Union’s Horizon 2020 research and innovation programme for the CoCa project under grant agreement No. 667302 and NIMH grants R01MH101519-01A1 and R01MH101519-01A1S1. This report reflects only the views of the authors and the commission bears no responsibility for any uses made of the information contained in the report.

